# Sex moderates apolipoprotein E ε4 effects on sleep expression and memory retention

**DOI:** 10.64898/2026.04.16.26351049

**Authors:** Negin Sattari, Abhishek Dave, Ivy Y. Chen, Kitty K. Lui, Miranda G. Chappel-Farley, Destiny E. Berisha, Kate E. Sprecher, Brady A. Riedner, Stephanie Jones, Barbara B. Bendlin, Bryce A. Mander, Ruth M. Benca

**Author notes:** **Corresponding authors:** Negin Sattari, PhD, Department of Psychiatry and Human Behavior, University of California, Irvine, Irvine,CA, United States, Bryce A. Mander, PhD, Department of Psychiatry and Human Behavior, University of California, Irvine, Irvine,CA, United States, Ruth M. Benca, MD, PhD, Department of Psychiatry and Behavioral Medicine, Wake Forest University School of Medicine, Winston-Salem, NC, United States. This study was not a clinical trial.

## Abstract

**Introduction:** Sleep-dependent memory consolidation differs by sex and maybe disrupted by Alzheimer’s disease (AD) risk. Whether sex moderates associations between apolipoprotein E ε4 (*APOE ε4*) status, non-rapid eye movement (NREM) sleep, and memory remains unclear.

**Methods:** Eighty cognitively unimpaired older adults completed a word-pair memory task with encoding and immediate testing occurring prior to overnight polysomnography with high-density electroencephalography (hdEEG) and delayed recall occurring after sleep. Sleep-memory associations were examined as a function of sex and *APOE ε4* status.

**Results:** In this sample, a sex×*APOE ε4* interaction was associated with overnight memory retention, with female carriers exhibiting less overnight forgetting than female non-carriers and male ε4 carriers. NREM sleep differed by sex and *APOE ε4* status and was associated with memory retention in *ε4* carriers.

**Discussion:** These findings indicate sex-specific, sleep-dependent memory mechanisms associated with genetic AD risk, highlighting sleep as a potential early target for intervention, pending replication in larger samples.

## 1. Background

Alzheimer’s disease (AD) is the most common cause of dementia and a major global health concern, with cases increasing as the population ages.^1^ The apolipoprotein E (*APOE*) *ε4* allele is a well-established and non-modifiable genetic risk factor for late-onset sporadic AD, associated with earlier diseaseAD onset, greater amyloid^2^ and tau burden^3^, and accelerated cognitive decline relative to *ε4* non-carriers.^4^ However, the biological pathways through which *APOE ε4* contributes to AD risk, particularly in asymptomatic individuals, remain incompletely understood. Notably, the impact of *APOE ε4* on AD risk and progression appears to differ by sex, another non-modifiable risk factor.^5,6^ Women not only experience AD at higher rates than men but may exhibit greater vulnerability to the pathological effects of the *APOE ε4* allele.^5,6^ Epidemiological studies suggest that female *ε4*-carriers show faster cognitive decline, steeper trajectories of brain atrophy, and greater tau accumulation^7^ compared to male carriers.^8,9^ Yet, despite this heightened risk, the mechanisms underlying sex-dependent effects of *APOE ε4* remain unclear.

Several studies indicate that sleep influencescognitive health and may contribute to AD pathogenesis.^10,11,12^ Non-rapid eye movement (NREM) sleep plays an important role in the consolidation of newly acquired information into long-term memory.^13,14^ Specific neural oscillations during NREM sleep, particularly sleep spindles and slow waves, are thought to support this process by facilitating hippocampal-cortical communication.^13,15,16^ Disruptions in these oscillations have been reported early in the course of AD and have been associated with impaired memory consolidation.^17,18^ Sleep is increasingly recognized as a potential modifiable risk factor for AD.^10,12^ In a seminal study,^19^ Prinz et al. (1982) demonstrated that individuals with AD exhibit alterations in sleep architecture, including reduced slow wave sleep and REM sleep and increased nighttime wakefulness.^19^ These changes differed between men and women, although the study did not formally assess whether AD-related sleep alterations interacted with sex beyond normative aging effects.

More recently, Lim et al. (2013) found that objectively measured sleep fragmentation, quantified via wrist actigraphy as an index of movement-related arousals and short immobile bouts, was associated with faster cognitive decline and increased AD incidence in older adults. In a prospective cohort, greater sleep fragmentation, measured with wrist actigraphy, predicted steeper declines in global cognition and higher AD risk over time.^20^ In a separate study, Lim et al. further showed that sleep fragmentation modified *APOE ε4* effects on AD risk and neurofibrillary tangle burden, such that *ε4* carriers with more fragmented sleep exhibited the highest pathological and clinical risk, whereas more consolidated sleep attenuated *ε4-*related vulnerability.^21^

Beyond their role in memory consolidation, NREM sleep oscillations may be implicated in AD pathophysiology.^22,23^ In mice, sleep enhances CSF–interstitial fluid exchange and promotes clearance of metabolites, including amyloid-β.^24^ In humans, NREM slow waves are temporally coupled to large-scale cerebrospinal fluid oscillations, suggesting a potential physiological mechanism linking sleep to brain clearance processes.^25^ Beyond clearance-related pathways, NREM oscillations, particularly the coordinated coupling of slow oscillations and sleep spindles, support synaptic plasticity and hippocampal–cortical communication.^15,16^ These plasticity-related processes are critical for systems level memory consolidation^14^ and apear disrupted early in AD.^17^

Accordingly, alterations in these oscillatory features may represent early physiological indicators of AD vulnerability.^17^ Given evidence that *APOE ε4* carriers show heightened susceptibility to synaptic and tau-related pathology,^26^ suggesting that disruptions in sleep-dependent plasticity mechanisms may be particularly relevant in genetically at-risk individuals.^11,26^ Together, these links between sleep physiology, memory consolidation, and AD-related pathology, suggest that sleep may represents a compelling candidate mechanism through which genetic and biological risk factors interact to influence early cognitive vulnerability.^18,27^

Genetic risk factors may modify the relationship between sleep neurophysiology and memory. *APOE ε4*, the strongest genetic risk factor for late-onset AD,^28^ has been associated with alterations in sleep physiology and memory function even in cognitively unimpaired adults, suggesting that genetic risk may modulate sleep-memory relationships. Although *APOE ε4*-related alterations in sleep architecture have been reported in large population-based cohorts, including increased N3 sleep duration among *ε4* homozygous men,^29^ less is known about how *APOE ε4* interacts with biological sex to affect sleep physiology and sleep-memory relationships in cognitively unimpaired older adults.^11^ Importantly, most prior work has either collapsed across sex or treated sex as a covariate, rather than testing sex as a biological moderator of sleep-memory relationships. As a result, potential sex-specific pathways linking genetic risk, sleep neurophysiology, and memory consolidation remain poorly understood. This gap is particularly consequential given that women have greater *APOE ε4*-related AD risk^5^ and that sex-specific relationships between sleep neurophysiology and memory consolidation have been previously observed.^30,31^ Importantly, much of the existing literature examining interactions between *APOE ε4* and sleep neurophysiology has relied on samples that were predominantly male, limiting insight into sex-specific sleep-genetic pathways.^11,32^ In contrast to *ε4* homozygotes, the majority of *ε4* carriers in the general population are heterozygous (ε3/ε4),^4^ underscoring the importance of studying more representative samples.

Women, despite bearing a disproportionate lifetime risk for AD, also exhibit higher prevalence of sleep disturbances in midlife and older adulthood, including insomnia symptoms and sleep fragmentation.^33^ Despite these differences, women remain underrepresented in mechanistic studies of sleep neurophysiology.^34^ Converging evidence suggests that sleep-dependent memory consolidation differs by biological sex.^35^ Across adulthood, women show differences in NREM EEG physiology relative to men, including higher slow wave activity and variation in sleep spindle characteristics. ^31,36^ Given the established role of NREM oscillations in memory consolidation,^15,16^ sex-related differences in these oscillatory dynamics may have functional implications for overnight episodic memory. Importantly, prior studies have reported sex differences in how NREM microstructure relates to sleep dependent memory improvement, with differential relationships between NREM oscillatory dynamics and sleep-dependent memory performance across men and women.^35^ Together, these findings suggest that NREM oscillatory physiology may differentially relate to overnight episodic memory in women, providing a biologically grounded rationale for testing sex as a moderator of *APOE*-related sleep-memory relationships.

Emerging evidence suggests that heterozygous *ε4* carriage may confer subtler and potentially sex-dependent neurophysiological effects that are not captured when genotype dose or sex is not explicitly modeled.^37,38,5,6,11^ As a result, analyses conducted in predominantly male samples may obscure meaningful variability in how genetic risk relates to sleep architecture, sleep microarchitecture, and sleep-dependent memory processes, particularly in women. These gaps motivated a sex-stratified investigation of how *APOE ε4* status relates to NREM sleep architecture, local sleep microarchitecture, and overnight episodic memory retention in older adults prior to cognitive impairment. This knowledge gap is particularly important in cognitively healthy older adults, where identifying early physiological markers could inform preventive interventions before clinical symptoms emerge. These mechanisms are likely to be most informative prior to the clinical manifestation of AD, when compensatory processes may still be engaged and modifiable.

In the present study, we examined whether sex moderates the relationship between *APOE ε4* status and sleep-dependent episodic memory retention in cognitively unimpaired older adults enriched for AD risk. Using high-density EEG and an overnight word-pair memory task, we further tested whether associations between NREM sleep architecture (i.e., N2 and N3 sleep stages), NREM EEG microarchitecture, and overnight memory retention vary as a function of sex and *APOE ε4* status. We hypothesized that sex would moderate associations among *APOE ε4* status, NREM sleep physiology, and overnight memory retention. Specifically, we predicted that sleep-memory relationships would vary by sex and *APOE ε4* status.

## 2. Methods

### 2.1. Participants

Eighty cognitively unimpaired older adults completed an overnight polysomnography (PSG) study with high-density electroencephalography (hdEEG; mean age=61.3±5.1 years; 49 female; 25 APOE ε4 carriers,14 female). This cohort was utilized to examine associations between sex, APOE genotype, and sleep expression. A subset of these participants (N=65; mean age=61.12±5.9 years; 40 female; 20 APOE ε4 carriers) additionally completed the word-pairs task to assess overnight memory retention. This subset was utilized to examine effects of sex, APOE genotype, and sleep expression on overnight sleep-dependent memory retention. Participants were recruited from the Wisconsin Alzheimer’s Disease Research Center (ADRC) clinical core. The cohort was enriched for AD risk^39^ characterized by parental history and APOE ε4 positivity (Table 1). The majority of APOE ε4 carriers in the present sample were heterozygous (ε3/ε4), with few ε4/ε4 individuals, precluding genotype dose-response analyses. Individuals with significant neurological, psychiatric, or medical conditions that could affect cognition or sleep were excluded. Recruitment was not stratified or targeted based on APOE genotype, and APOE status was determined following enrollment and was not used as a selection criterion for participation. Additionally, use of medications known to influence sleep or EEG (e.g., antipsychotics, sedatives, SSRIs) was exclusionary. Cognitive status was confirmed through comprehensive neuropsychological testing, including the National Alzheimer’s Coordinating Center Uniform Data Set (UDS)= and additional validated assessments.^41,42^ All participants provided written informed consent, and study procedures were approved by the Institutional Review Board at the University of Wisconsin-Madison.

**Table 1.**
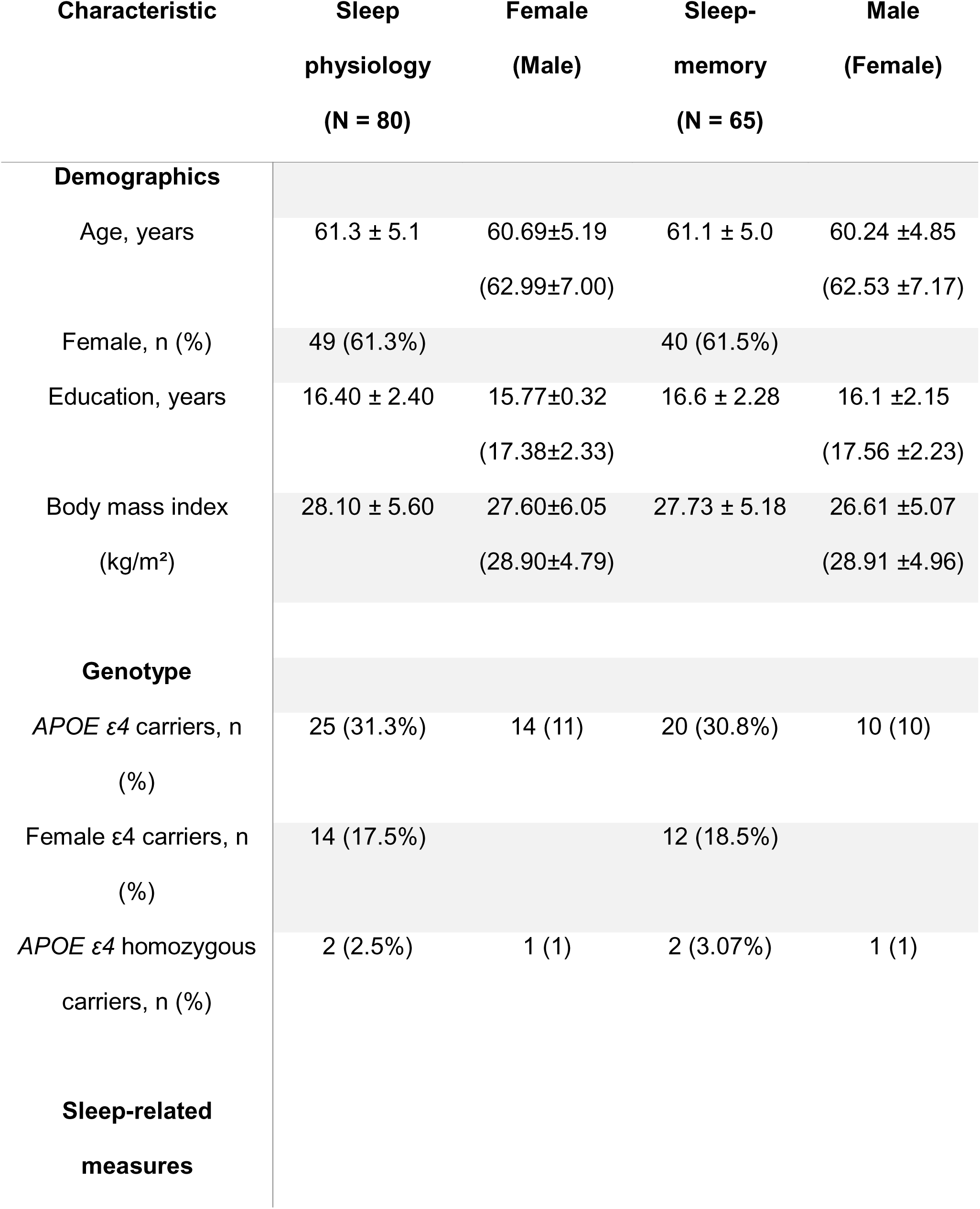
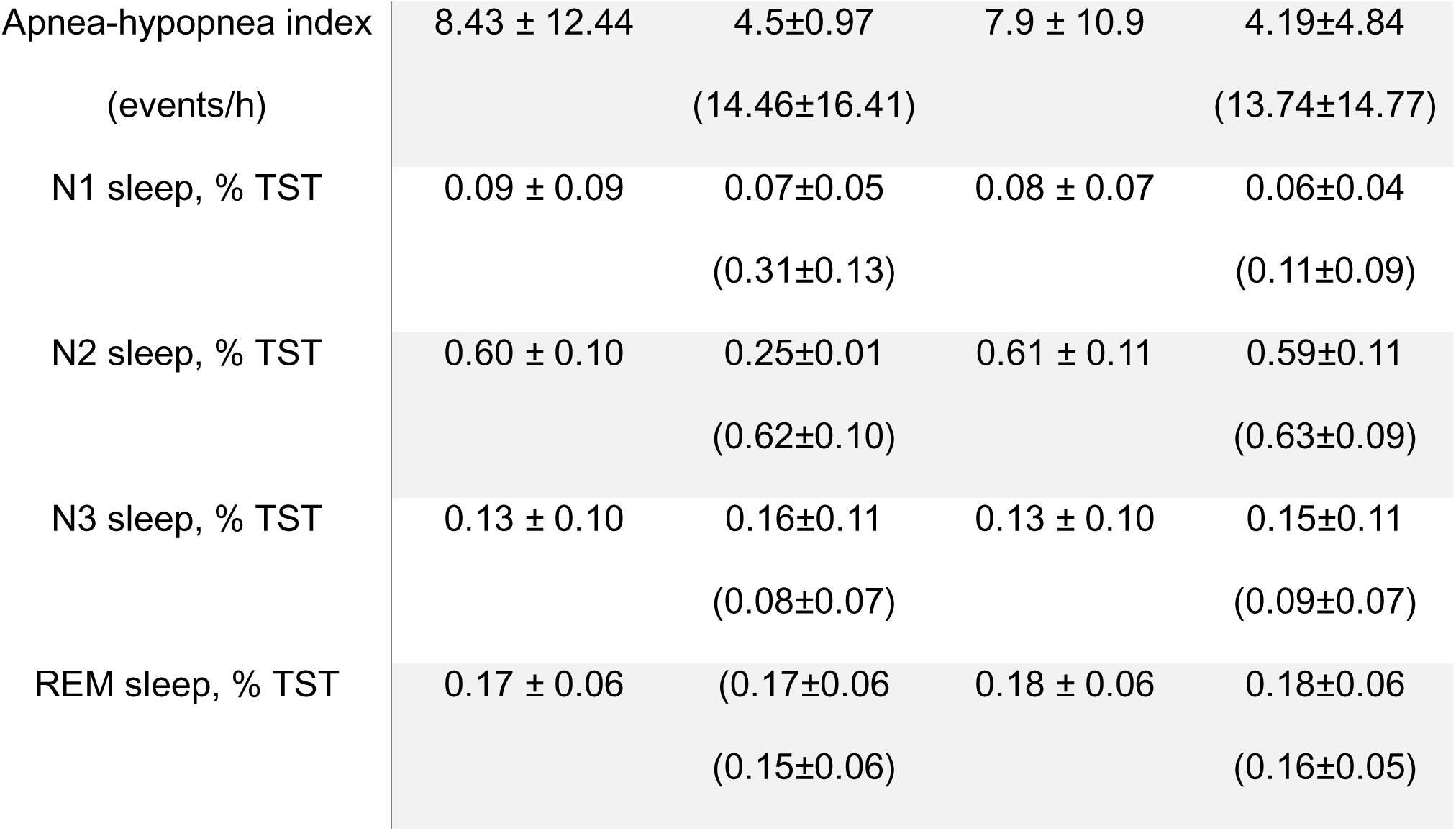
Participant characteristics. Note: The sleep physiology sample (N = 80) includes all pa rticipants with valid overnight EEG data. The sleep-memory sample (N = 65) includes participants with complete overnight memory data. Demographic and clinical characteristics were comparable across samples. All reported means are presented with their corresponding standard deviations (mean± SD).

### 2.2. Study Protocol

Participants completed an in-laboratory evening session of memory encoding followed by an immediate recall test, overnight polysomnography with hdEEG, and a delayed recall test the following morning to assess sleep-dependent memory consolidation. APOE genotype was determined from whole blood samples using DNA extraction followed by competitive allele-specific PCR-based KASP genotyping for rs429358 and rs7412, as previously described.^43^ The final analytic sample consisted of 80 participants with PSG data and 65 participants with memory data at both immediate and delayed recall testing sessions.

### 2.3. Polysomnography (PSG) and EEG Acquisition

Overnight sleep was assessed using standard clinical polysomnography (PSG) and concurrently recorded hdEEG.^44^ PSG data were collected using a customized Alice 5 System (Philips Respironics, Murrysville, PA, USA) and included electrooculogram (EOG), submental electromyogram (EMG), electrocardiogram (ECG), respiratory inductance plethysmography, pulse oximetry, and body position monitoring. Simultaneously, 256-channel hdEEG was recorded using a NetAmps 300 amplifier and NetStation software (Electrical Geodesics Inc., EGI), with vertex referencing. EEG signals were digitized at 500 Hz. Sleep staging was performed by a registered polysomnographic technologist according to American Academy of Sleep Medicine (AASM) criteria^45,46^ using six standard 10-20 EEG derivations (F3, F4, C3, C4, O1, O2) extracted from the hdEEG montage and re-referenced to the contralateral mastoid. Sleep scoring was conducted using Alice Sleepware and reviewed in full by a board-certified sleep physician (co-author R.M.B.), including verification of clinical sleep events. Standard sleep architecture measures were derived, including total recording time (TRT), total sleep time (TST), sleep latency (SL), sleep efficiency (SE), wake after sleep onset (WASO), and percentage of time spent in each sleep stage (N1, N2, N3, REM). In addition, total sleep apnea-hypopnea index (AHI; number of apneas and hypopneas per hour) which is a clinical measure characterizing sleep-disordered breathing was also derived. AHI is a traditional measure for Obstructive Sleep Apnea (OSA) diagnosis, consistent with American Academy of Sleep Medicine (AASM) clinical criteria defining at least mild OSA as an AHI index≥ 5 events per hour. ^47,48^

### 2.4. EEG Spectral Analysis

Preprocessing and spectral analyses were conducted using custom MATLAB scripts with EEGLAB toolboxes (http://sccn.ucsd.edu/eeglab). As previously described,^44^ EEG data were notch-filtered in the 59.5-60.5 Hz range to remove line noise and then bandpass-filtered between 0.3-55 Hz using a 5500-point Hamming-windowed sinc Finite Impulse Response filter for both operations. The 50 outermost electrodes were excluded from further analyses due to recurring physiological artifacts at these locations across participants. The remaining 206 electrodes were average-referenced and manually inspected for persistent, long-duration noise to remove compromised channels. Data corresponding to NREM sleep stages (N2 and N3) were then extracted from these electrodes and concatenated into separate matrices for subsequent analyses. A three-stage semi-automated artifact rejection procedure was subsequently applied to eliminate short durations of widespread noise and consistently noisy EEG derivations.^49–51^ Epochs with arousals were first removed. Next, an automated 99.8th-percentile threshold on broadband power was applied to exclude high-amplitude artifacts. Residual short-duration artifacts and noisy electrodes were then visually rejected. Finally, deleted channels were reconstructed using spherical spline interpolation, yielding clean NREM EEG data for all 80 subjects.^52,53^ Next, spectral analyses were performed using a multitaper Fast Fourier Transform on cleaned EEG data to estimate absolute and relative power from 0.5-40 Hz. Eleven Slepian tapers (bandwidth = 0.25 Hz) were applied with zero padding and 30 s windows sliding every 5 s. The resulting time-frequency matrix (μV²/Hz) was integrated and normalized to total power within each channel to derive relative spectral power (unitless ratio), indicating the proportion of power within each frequency band. Median spectral power was computed across NREM epochs for each of the 1,294 frequency bins, then averaged within canonical frequency bands: slow wave activity: 0.5-1.5, delta:1.5-4.5, theta: 4.5-7.5Hz, alpha: 7.5-11Hz, slow sigma: 11-13Hz, fast sigma: 13-16Hz, beta: 16-28Hz, and gamma: 28-40Hz). Spectral analyses were performed using the Chronux v2.12 toolbox (http://chronux.org) in MATLAB. Analyses focused a priori on delta and sigma frequency bands because of their relevance for slow waves and sleep spindles that are known to support memory consolidation, and to be impacted by sex, aging, and AD.^13–15^

### 2.5. Word-Pairs Memory Task

Overnight episodic memory retention was assessed using a validated word-pair task that is sensitive to sleep-dependent memory consolidation in both young adults and individuals with mild cognitive impairment (MCI).^54–57^ The task was administered using E-Prime software (Psychology Software Tools, Pittsburgh, PA) and consisted of three phases: encoding, immediate recall, and delayed recall. The encoding phase occurred in the evening prior to sleep onset. During this phase, participants were presented with 88 semantically unrelated word-pairs, each displayed sequentially for four seconds with one word positioned above the other at the center of a computer monitor. Participants were instructed to learn the word-pairs for later recall. To minimize primacy and recency effects, the first and last four word-pairs were excluded from subsequent recall phases. Immediately following encoding, participants completed an immediate recall phase in which one word from each pair was presented individually on the screen, and participants were instructed to type the corresponding word. Responses were self-paced, and word-pair order was randomized across participants and task phases. Participants were seated with the experimenter out of view to minimize feedback-related bias. After completion of the immediate recall phase, participants were shown feedback indicating their percentage accuracy. Encoding and immediate recall phases were repeated until participants achieved a minimum of 60% accuracy; performance from the final immediate recall phase was used for analysis. The delayed recall phase was administered the following morning, approximately one hour after sleep offset, and was completed only once without performance feedback. Accuracy during both immediate and delayed recall phases was calculated as the proportion of correctly recalled word-pairs. Responses containing clear typographical errors (e.g., forg instead of frog) were corrected offline prior to analysis. Overnight memory retention was quantified as the proportional change in recall performance from immediate to delayed testing, calculated as (Delayed Recall-Immediate Recall)/Immediate Recall. To address non-normality in the distribution of this variable, a cube-root transformation was applied to this proportional change prior to statistical analyses.

### 2.6. Data Reduction

Participants were included in sleep-dependent memory analyses if both immediate and delayed recall data were available. Fifteen participants were excluded from memory analyses due to missing memory data (n = 11) or identification as statistical outliers (n = 4). Outliers were defined as a priori as values exceeding ±3 standard deviations from the mean of the overnight memory retention metric. Participants failing to meet the≥60% performance criterion at immediate recall were excluded prior to these analyses. To assess potential selection bias, excluded from the memory analysis participants were compared to included participants on age, sex, and APOE ε4 status using independent samples t-tests. No significant differences were observed (all p > .21).

### 2.7. Statistical Analysis

Analyses followed a hierarchical approach, first testing sex×*APOE ε4* interactions, followed by stratified analyses only when interaction terms reached statistical significance. A univariate ANCOVA was conducted to examine sleep-dependent memory consolidation, with overnight memory retention as the dependent variable with sex and *APOE ε4* status as fixed factors. Age, apnea-hypopnea index (AHI), and body mass index (BMI) were included as covariates given their established associations with sleep physiology and cognitive performance in older adults.^10^ These analyses included participants with available immediate and delayed recall data (n = 65). A multivariate ANCOVA was conducted to examine effects of sex and *APOE ε4* on sleep architecture, with percentage of time spent in each sleep stage as dependent variables, sex and *APOE ε4* status as fixed factors, and age, education, and AHI as covariates. These analyses included all participants with PSG data (n = 80). Age, years of education, and AHI were included as covariates given their known associations with sleep architecture and cognitive performance and because they did not differ systematically across sex or *APOE ε4* groups. Significant interactions were followed by planned post hoc comparisons using least significant difference tests to examine simple effects within sex and *APOE ε4* groups. Post hoc analyses were conducted only when omnibus interaction terms reached statistical significance. Pearson’s correlations were employed to examine sex- and *APOE ε4*-related associations between memory retention and sleep architecture measures.

For sleep EEG spectral power analyses assessing associations with overnight changes in word-pair memory performance across groups, Pearson’s r values were first computed at each electrode across the scalp. The Threshold-Free Cluster Enhancement (TFCE) procedure (5,000 permutations; α = 0.05; extent = 0.5; height = 2.0) was then applied to correct for multiple comparisons across topography.^58–60^ Absolute spectral power from electrodes that remained significant after TFCE correction was extracted, averaged, and entered into regression models predicting overnight memory change, adjusting for age, education, body mass index, and AHI to account for potential confounding influences on sleep physiology and cognition. To evaluate group differences in spectral power, independent-samples t-tests were conducted at each electrode, followed by TFCE correction (5,000 permutations) for multiple comparisons across topography. TFCE correction for multiple comparisons analysis was implemented using a custom MATLAB (R2017b; MathWorks, Natick, MA, USA) derived from previously described nonparametric permutation methods.^58–60^ EEG preprocessing and data handling relied on EEGLAB.^61^

All of the analyses were conducted using SPSS (Version 28.0.0.0) (IBM SPSS Statistics, Inc., Chicago, IL).

## 3. Results

### 3.0. Sample Characteristics and Data Completeness

Of the 80 participants who completed overnight polysomnography and 65 had complete the memory task. To assess potential selection bias, participants included in memory analyses were compared to those excluded on age, sex, and APOE ε4 status. No significant differences were observed between included and excluded participants (all p values > .21), indicating that the analytic sample was not systematically biased.

### 3.1. Sex, *APOE ε4* Status, and Sleep-dependent Memory Retention

Sleep-dependent memory analyses were conducted in a final sample of 65 participants. We first examined if the AD risk factors of sex and *APOE ε4* status impacted overnight sleep-dependent memory retention. A univariate ANCOVA revealed a significant sex × *APOE ε4* interaction on overnight memory retention, F(1, 56) =7.36, p =.009, partial η² =.116, after adjusting for age, AHI, and BMI. The overall model showed a trend toward significance, F(6,56) =2.02, p =.079. Post hoc comparisons confirmed no difference in overnight memory retention between *APOE ε4* carriers and non-carriers among male participants [t(23) =.91, p=.37], Figure 1. Among women however, we found a significantly less overnight forgetting among *APOE ε4* carriers compared to non-carriers [t(38) =2.52, p=.04]. Furthermore, memory retention among male *APOE ε4* carriers was worse compared to female carriers [t(18) =-2.11, p=.05]. These results suggest that overnight sleep-dependent memory retention in cognitively unimpaired older adults may vary as a function ofboth sex and *APOE ε4* status, such that sex differences in verbal memory retention appear more pronounced among *APOE ε4* carriers. Given the modest sample size, these findings should be interpreted as preliminary.

**Figure 1:**
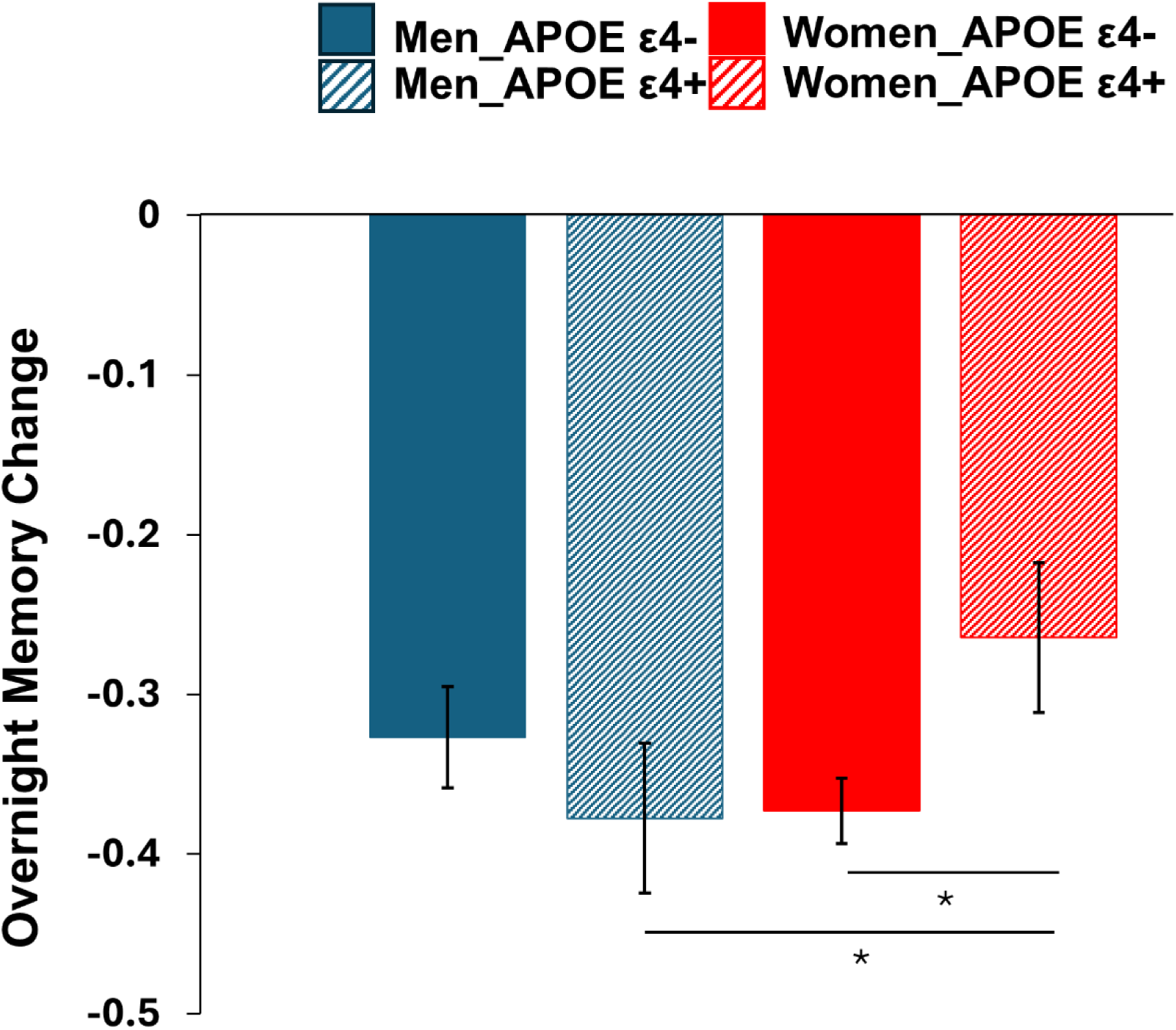
Sleep-dependent memory retention as a function of sex and *APOE ε4* status. Solid bars represent *APOE ε4* non-carriers and hatched bars represent *APOE ε4* carriers. Blue bars denote men and red bars denote women. Bars show group means, and error bars represent ± SEM. Overnight memory retention was calculated as proportional change from immediate to delayed recall [(Delayed Recall - Immediate Recall) / Immediate Recall]. Statistical analyses were performed on cube-root-transformed proportional change scores to address non-normality. Asterisks (*) indicate significant group differences (p < .05).

### 3.2. Sex, *APOE ε4* Status, and Sleep Architecture

We next examined whether sex and *APOE ε4* status were associated with sleep architecture measures, including percentage of time spent in N1, N2, N3, and REM sleep stages. Multivariate ANCOVA was conducted to predict sleep stage variables, with sex and *APOE ε4* status as between-subjects factors and age, years of education, and AHI as covariates. The overall models were significant for N1, [F(6, 72)=5.93, p<.001, partial η² =.33], N2, [F(6, 72)= 2.29, p=.044, partial η²=.16], and N3 sleep stages, [F(6, 72)=3.02, p=.011, partial η²=.20], but not for REM sleep, [F(6, 72)=1.58, p=.17, partial η²=.12]. For N1 sleep, older age at sleep study was associated with increased N1 sleep percentage, [F(1, 72)=15.41, p<.001, partial η²=.18]. No significant main effects of sex or *APOE ε4* status were observed for N1 sleep, although the sex×*APOE ε4* interaction approached significance, [F(1, 72)=3.80, p=.055, partial η²=.05]. For N2 sleep, a significant sex×*APOE ε4* interaction was observed, [F(1, 72)=6.97, p=.010, partial η² =.09]. Post hoc analyses were conducted to characterize the interaction, Figure 2. These analyses indicated that *APOE ε4* carrier status was associated with greater N2 sleep in men but not in women (p=.03). No significant main effects of sex or *APOE ε4* status were detected. For N3 sleep, a significant main effect of sex was observed [F(1, 72)=7.25, p=.009, partial η²=.09] indicating sex differences in slow wave sleep percentage. No significant main effect of *APOE ε4* status or sex×*APOE ε4* interaction was detected for N3 sleep. Together, these findings indicate that sex and *APOE ε4* status are associated with distinct patterns of NREM sleep architecture, including a sex×*APOE ε4* interaction for N2 sleep duration and a main effect of sex on N3 sleep duration. No significant main effects or interactions were observed for REM sleep (all p values>.05).

**Figure 2:**
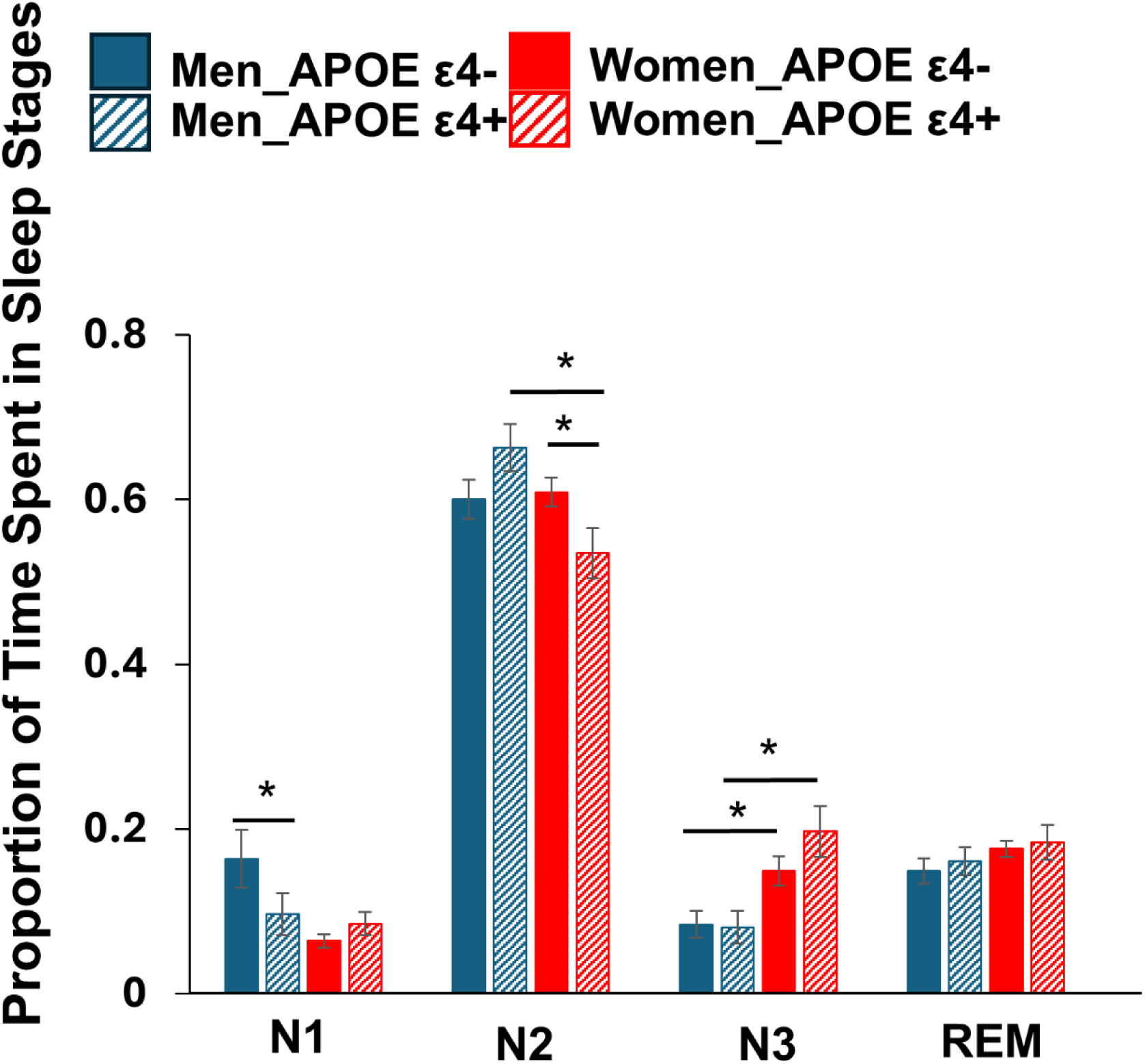
Sleep architecture measures stratified by sex and *APOE ε4* status. Percentage of total sleep time (TST0 spent in N1, N2, N3, and REM sleep is shown for men (blue) and women (red), with solid bars representing *APOE ε4* non-carriers and hatched bars representing *APOE ε4* carriers. Bars depict group means, and error bars represent ± SEM. Asterisks (*) indicate significant group differences (p < .05).

We next examined whether sex and *APOE ε4* status were associated with measures of sleep continuity, including WASO and SE. Separate univariate ANCOVAs were conducted for WASO and SE, with sex and *APOE ε4* status as between-subjects factors and age, years of education, and AHI as covariates. The overall model for WASO did not approach statistical significance, [F(6, 72)=2.06, p=.069, partial η²=.15]. No significant sex×*APOE ε4* interaction was observed for WASO, [F(1, 72)=.02, p=.90, partial η²<.001]. A significant main effect of age was detected, [F(1, 72)=8.94, p=.004, partial η²=.11], such that older age was associated with increased WASO. No significant main effects of sex or *APOE ε4* status were observed (all ps>.05). The overall model for sleep efficiency was also not statistically significant, [F(6, 72)=1.46, p=.20, partial η²=.11]. No significant sex×*APOE ε4* interaction was observed, [F(1, 72)=.15, p=.705, partial η²=.002]. A significant main effect of age was observed, [F(1, 72)=7.06, p=.01, partial η²=.09], indicating lower sleep efficiency with increasing age. No significant main effects of sex or *APOE ε4* status were detected (all p values>.05). These findings indicate that the effects of sex, *APOE ε4* status, and their interaction on sleep architecture may be independent of age effects on sleep fragmentation.

### 3.3. The Role of Sleep Architecture in Sleep-dependent Memory

Given observed effects of sex and *APOE ε4* status on sleep architecture and overnight memory retention, we next examined associations between sleep architecture and overnight memory retention across sex and *APOE ε4* status using Pearson correlations. When examining these associations in men and women separately while collapsing across *APOE ε4* status, N2 sleep duration was positively associated with overnight memory retention in men (r=.399, p=.048), whereas no significant associations between sleep stage duration and overnight memory retention were observed in women (all p values>.09). When analyses were further stratified by *APOE ε4* status, the N2-memory association in men was driven by *APOE ε4* carriers, with greater N2 sleep duration being associated with better overnight memory retention (r =.67, p=.03), Figure 3. In contrast, greater REM sleep duration was associated with poorer overnight memory retention (r =−.78, p=.007) in male *APOE ε4* carriers, indicating greater overnight forgetting with increasing REM sleep. However, among female *APOE ε4* carriers, greater slow wave sleep (SWS; N3) duration was positively associated with overnight memory retention (r=.71, p=.02), Figure 3. None of these associations in men and women were significant among *APOE ε4* non-carriers (all p values>.35). Collectively, these findings suggest that sex moderates sleep-memory relationships, particularly among *APOE ε4* carriers, with N2 preferentially associated with memory retention in men and N3 preferentially associated with memory retention in women. Notably, these associations were selective to *APOE ε4* carriers and were absent in non-carriers, underscoring the influence of genetic AD risk factors on sleep-memory relationships. Given the limited sample size, absence of convergent REM effects across analyses, and the interdependence of sleep stages, REM-related findings should be interpreted cautiously, as associations with REM sleep may reflect reciprocal reductions in NREM sleep rather than REM-specific mechanisms.

**Figure 3:**
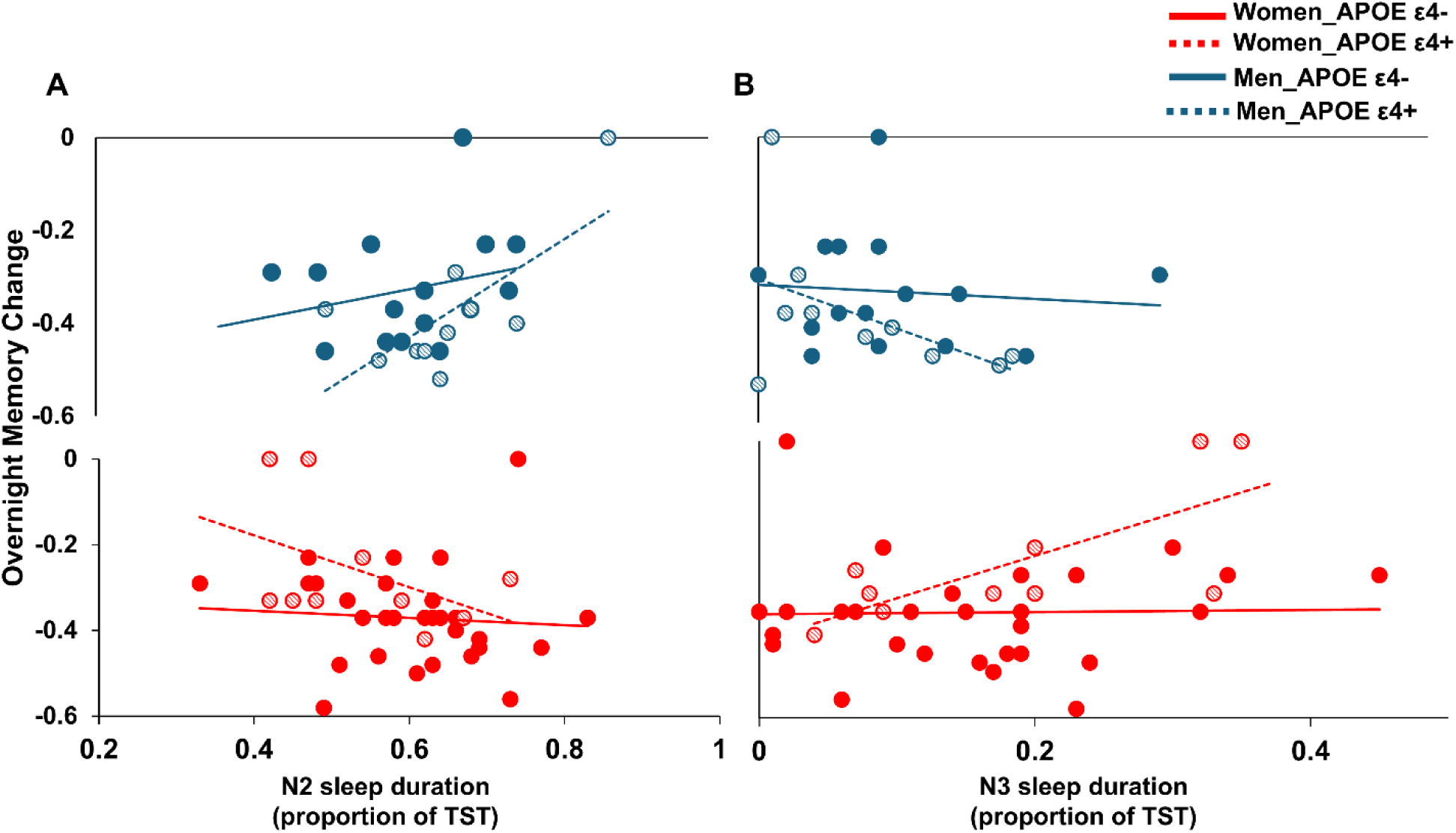
Associations between NREM sleep stages (N2, N3) and overnight sleep-dependent memory retention stratified by sex and *APOE ε4* status. (A) Relationship between N2 sleep duration (proportion of total sleep time, TST) and overnight memory change. (B) Relationship between N3 sleep duration (proportion of TST) and overnight memory change. Blue symbols denote men (top panel) and red symbols denote women (bottom panel). Solid lines represent *APOE ε4* non-carriers and dashed lines represent *APOE ε4* carriers. Points represent individual participants. Overnight memory retention was calculated as proportional change from immediate to delayed recall [(Delayed Recall - Immediate Recall) / Immediate Recall]. Statistical analyses were performed on cube-root-transformed proportional change scores. Regression lines are shown for visualization, with corresponding R² values: Men ε4− (N2=.08; N3=.01), Men ε4+ (N2=.50*; N3=.26), Women ε4− (N2=.007; N3=.0009), Women ε4+ (N2=.20; N3=.53*). Asterisks (*) indicate statistically significant associations (p<.05).

### 3.4. Sex, *APOE ε4* Status, and NREM Sleep Neurophysiology

We next examined whether local expression of NREM sleep microarchitecture differed as a function of sex and *APOE ε4* status. When comparing men and women, significant sex differences in NREM spectral power were observed. Relative to men, women demonstrated greater global slow oscillation (SO) power (.5-1.5 Hz; p=.01; Figure 4-A) and delta band power (1.5-4.5 Hz; p=.01; Figure 4-B) compared to men, with electrodes over central and parietal regions surviving TFCE correction. Women also exhibited significantly higher slow sigma power over central electrodes following TFCE correction for multiple comparisons (11-13 Hz; p=.02; Figure 4-C). Next, we examined whether *APOE ε4* status modulated NREM sleep microarchitecture within each sex. Following TFCE correction for multiple comparisons, *APOE ε4*-related differences in NREM spectral power were observed only among women, with no comparable effects detected in men. Specifically, female *APOE ε4* carriers exhibited significantly higher frontal slow sigma power compared to female non-carriers following TFCE correction, Figure 4-D. No significant differences in NREM spectral power were detected between *APOE ε4* carriers and non-carriers among men after correction for multiple comparisons, although smaller effects may not have been detectable given the sample size.

**Figure 4:**
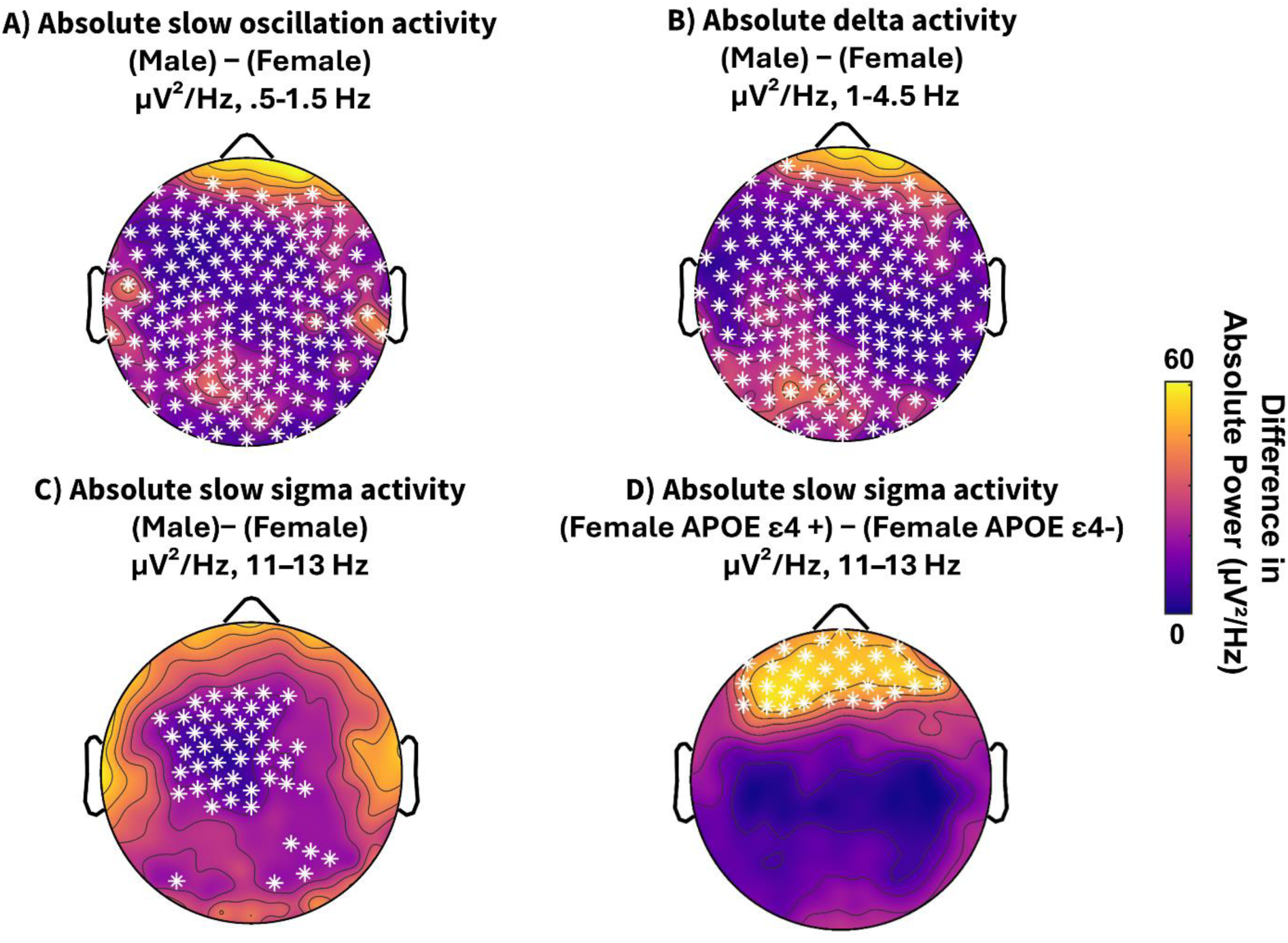
Sex- and genotype-related differences in NREM spectral power. Topographic maps depict between group differences in NREM spectral power (µV²/Hz). Panels A-C show male-female differences in (A) slow oscillation power (.5-1.5 Hz), (B) delta power (1.5-4.5 Hz), and (C) slow sigma power (11-13 Hz), computed as male minus female. Panel D shows slow sigma power differences (11-13 Hz) between female *APOE ε4* carriers and non-carriers, computed as carriers minus non-carriers. Warmer colors indicate greater slow sigma power in *ε4* carriers relative to non-carriers, whereas cooler colors indicate the opposite pattern. Asterisks denote electrodes surviving Threshold-Free Cluster Enhancement (TFCE) correction for multiple comparisons (p < .05). TFCE integrates spatial extent and statistical magnitude into a single nonparametric test; no additional cluster-level analysis was performed. Color shading is shown for visualization purposes only; statistical inference is based solely on TFCE-corrected electrode-level statistics.

### 3.5. Sleep Neurophysiology and Sleep-Dependent Memory

Lastly, given the detected associations with sleep architecture and overnight memory retention, we examined how local expression of NREM sleep microarchitecture was associated with overnight memory retention stratified by sex and *APOE ε4* status. When examined separately in men and women, while collapsing across *APOE ε4* status, significant associations between NREM sleep microarchitecture and overnight memory retention were observed only in women. Specifically, greater slow sigma and fast sigma power during NREM sleep were positively associated with better overnight memory retention in women (all p values<.05). Analyses guided by our *a priori* focus on NREM delta and sigma power revealed no significant associations with overnight memory retention in men (all p values>.10). We next examined whether these associations differed as a function of *APOE ε4* status. Among female *APOE ε4* carriers, there were positive associations between overnight memory retention and slow sigma [r=.79, p=.009], fast sigma [r=.72, p=.02], and delta [r=.73, p=.01] activity, Figure 5. In contrast, no significant associations between NREM sleep microarchitecture and overnight memory retention were observed among *APOE ε4* non-carriers or among men, irrespective of *APOE ε4* status. These findings suggest that NREM sleep microarchitecture is particularly relevant for memory consolidation in women with increased genetic risk for AD. However, given the modest subgroup sample sizes, these correlations should be interpreted as descriptive and hypothesis-generating rather than definitive.

**Figure 5.**
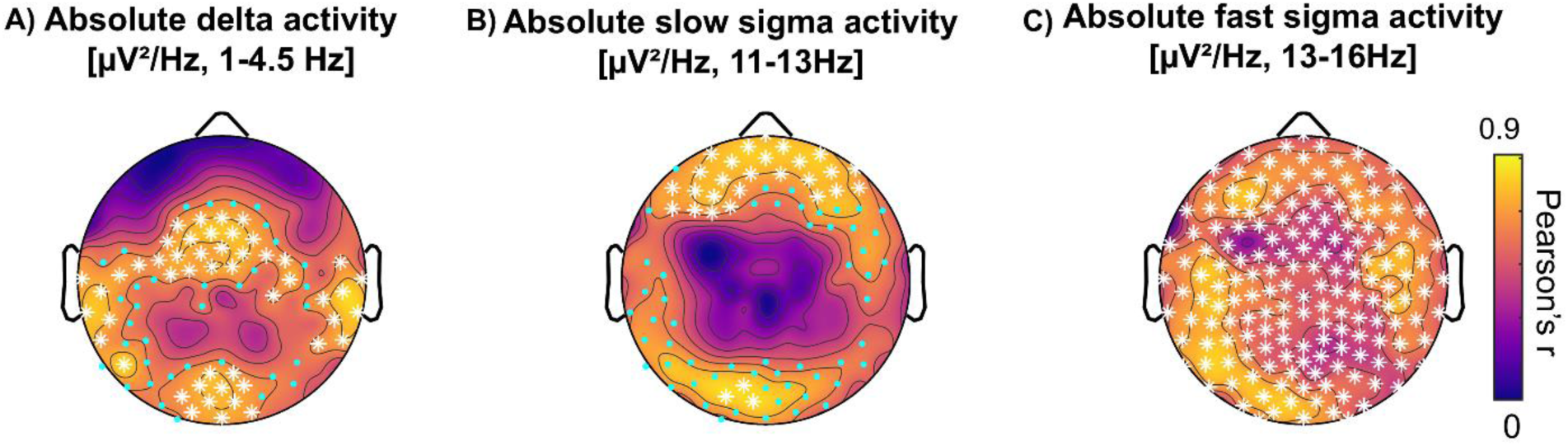
Topographic associations between NREM spectral power and overnight memory change in female *APOE ε4* carriers. Topographic maps illustrate Pearson correlation coefficients (r) between overnight memory change and absolute NREM spectral power for (A) delta activity (1.5-4.5 Hz), (B) slow sigma activity (11-13 Hz), and (C) fast sigma activity (13-16 Hz). Associations are plotted for female *APOE ε4* carriers only, as no other groups showed significant associations. Warmer colors indicate stronger positive associations. Blue dots indicate electrodes reaching uncorrected p < .05 for visualization purposes only, whereas asterisks denote electrodes surviving TFCE correction (p < .05). Statistical inference is based solely on TFCE-corrected electrode-level results. Scalp orientation is shown with the nose at the top. Spectral power is expressed in μV²/Hz.

## 4. Discussion

The central contribution of this study is providing evidence that sex moderates the relationship between *APOE ε4* status, NREM sleep physiology, and sleep-dependent memory consolidation in cognitively unimpaired older adults. Using hdEEG and an overnight word-pair paradigm, we found that sex and *APOE ε4* status interacted to shape memory outcomes in this sample, such that female *ε4* carriers exhibited less overnight forgetting than female non-carriers and male carriers. At the level of sleep architecture, sex and *APOE ε4* status were associated with differences in NREM sleep architecture. Associations between NREM sleep and overnight memory retention appeared to differ by sex and were most evident among*ε4* carriers. At the microarchitectural level, sigma activity during NREM sleep showed the strongest associations with memory retention, particularly among female *ε4* carriers. Together, these findings suggest that the relationship between *APOE ε4* status and sleep-related memory processes differs by sex, underscoring the importance of considering sex as a biological variable when examining sleep and cognition. However, given the modest sample size, these findings should be interpreted as preliminary and warrant replication in larger cohorts.

### 4.1. Sex-Specific Effects of *APOE ε4* on Memory Retention

We found that *APOE ε4* carrier status was not associated with overnight memory retention in men, but in women, *ε4* carriers demonstrated less overnight forgetting than non-carriers. Moreover, male *ε4* carriers exhibited poorer memory retention than their female counterparts, suggesting that *APOE ε4*-related differences in sleep-dependent memory retention may vary by sex, rather than reflecting a uniformly deleterious effect.. Prior work indicates that *APOE ε4* effects can be age-dependent and pleiotropic, with younger or preclinical carriers sometimes showing preserved or even enhanced cognitive performance before later decline.^5,6,62–66^ This sex-specific pattern^67^ is particularly interesting because *ε4* carriers are typically considered at greater risk for cognitive decline. The enhanced sleep-memory associations observed here may reflect compensatory or adaptive processes in the context of genetic risk, consistent with cognitive reserve and neural compensation frameworks,^68,69^ or alternatively, differences in how sleep-dependent mechanisms support memory at this preclinical stage.^11,38,62^ Such compensatory processes may temporarily enhance performance in midlife or early older adulthood, before giving way to accelerated decline at later disease stages.^5,7,70^ These findings are consistent with a recent report showing that SWA may be more predictive of memory in older adults with greater amyloid-β burden,^71^ highlighting memory-relevant sleep oscillations as candidate mechanisms of cognitive reserve in adults at risk for AD. These interpretations remain speculative and require longitudinal validation to determine their relevance for cognitive decline.

### 4.2. Sex Differences in Sleep Architecture and Their Relation to *APOE ε4*

Our findings suggest sex-specific patterns in sleep architecture and sleep–memory relationships that vary by *APOE ε4* status. Sex and genotype interacted to influence N2 duration, and women spent a greater proportion of the night in N3 sleep overall. However, *APOE ε4* status was not associated with N3 duration and did not interact with sex. Sex differences in sleep-memory associations were most evident when analyses were stratified by *APOE ε4* status. Sleep-memory associations differed by sex among *ε4* carriers, with stage-specific patterns emerging in men and women. Although circulating sex hormone levels are substantially reduced in postmenopausal women, lifetime hormonal exposure and menopausal transition-related changes in sleep physiology may contribute to sex-specific differences in NREM oscillatory dynamics. ^34,72,73^ While the present study did not directly assess hormone levels, future work incorporating endocrine measures is needed. Notably, these associations were not observed among *APOE ε4* non-carriers, suggesting that genetic risk may modulates the functional relevance of specific sleep physiology for memory in a sex-dependent manner.^5,11,18,64^ Of note, these findings do not suggest that greater N3 sleep universally supports memory consolidation in older adults. Rather, consistent with prior work demonstrating an age-related decoupling between slow wave sleep and episodic memory, associations between N3 duration and memory retention in the present study were limited and context-dependent.^11,17,74^ This pattern aligns with evidence suggesting that increases in slow wave sleep in later life may reflect altered or potentially excessive synaptic downscaling rather than enhanced consolidation, particularly in the context of AD risk.^17,18,75^

These findings build prior population-based work linking *APOE ε4* status to alterations in slow wave sleep, including reports of increased N3 duration among ε4/ε4 homozygous men in the Osteoporotic Fractures in Men (MrOS) cohort.^29^ In contrast to those studies, the present sample was composed predominantly of *ε4* heterozygotes, with few *ε4* homozygotes, limiting our ability to examine genotype dose effects. Consistent with Tranah et al., we did not observe differences in N3 sleep duration between *ε4* heterozygote carriers and non-carriers among men.^29^ Importantly, by including women and by focusing on sleep-memory associations rather than sleep stage duration alone, our results suggest that in predominantly heterozygous samples, sex-specific effects of *APOE ε4* may be more apparent in the functional relationship between sleep stages and memory performance than in sleep architecture per se.

### 4.3. Sex and *APOE ε4* Interact to Influence Sleep EEG-Memory Associations

We observed positive associations between NREM sleep spindle-relevant slow and fast sigma power and overnight memory retention in female *APOE ε4* carriers. No such associations were observed in males or *ε4* non-carriers of either sex. Although these associations were robust at the descriptive level, their attenuation in covariate-adjusted models warrants caution in inferring specificity beyond shared variance with demographic and sleep-related factors and suggests they should be interpreted as hypothesis-generating. These patterns may reflect a sex specific pleiotropic effect of *APOE ε4*^76^ or a compensatory mechanism in the face of elevated AD risk.^5,11,68^ These findings align with prior work linking sleep spindles to memory consolidation and suggest that sex-specific neural mechanisms may modulate their functional relevance.^11,15,31,77^. Sigma band activity, which reflects sleep spindle dynamics, has been repeatedly implicated in hippocampal-cortical communication and synaptic plasticity.^15,16,77,78^ The specificity of sigma-memory associations to female *APOE ε4* carriers suggests that spindle related mechanisms may be particularly sensitive to the interaction between sex biology and genetic risk, and may represent a candidate early, sex-specific marker of AD vulnerability.^17,18,31^

Sleep stages are defined by their underlying oscillatory dynamics. These findings suggest that variability in slow waves and spindles, rather than stage labels alone, may more precisely index mechanisms linking sleep to *APOE*-related vulnerability. Framing sleep in terms of microstructural oscillatory activity may therefore offer greater mechanistic specificity than traditional stage-based classifications. Although effects were localized to canonical spindle-generating regions and may be detectable with conventional EEG, hdEEG provides greater spatial resolution and sensitivity to subtle regional variation, potentially improving detection of early microstructural alterations associated with genetic risk.

### 4.4. Implications and Future Directions

Together, these results highlight the need for a sex- and genotype-sensitive approach to studying mechanistic pathways that contribute to cognitive function and AD risk. In particular, the findings suggest that female *APOE ε4* carriers may exhibit stronger associations between sleep physiology and memory, possibly reflecting an early adaptive response that may diminish with advancing age or disease progression.^11,38,68,69^ Several studies have suggested a pleiotropic effect of *APOE* that depends on age and sex; understanding these interactions is expected to help clarify why women are at increased risk of dementia due to AD^79^ and may help inform the development of early interventions, such as sleep-based strategies, tailored by sex and genetic risk. These findings also raise the possibility that interventions targeting NREM oscillatory activity may have differential efficacy depending on sex and *APOE ε4* status, “highlighting the potential value of precision approaches to AD prevention.

An importantcontribution of this study is the identification of sex- and *APOE ε4*-specific associations between sleep microarchitecture and sleep-dependent memory retention. In women, particularly *ε4* carriers, overnight memory retention was most strongly associated with NREM sigma activity.^11,17,74^ These findings converge with evidence that sigma related activity plays a key role in memory consolidation in older adults^17,77^ and suggest that microarchitectural features of NREM sleep may represent more sensitive early indicators of AD risk than conventional sleep staging.^11,18^

### 4.5. Limitations

Several limitations warrant consideration. First, the modest sample size, particularly within sex-by-genotype subgroups, limited power to detect higher-order interactions and may constrain generalizability. Additionally, a subset of participants was excluded from memory analyses due to missing data or outlier criteria; however, included and excluded participants did not differ on key demographic or genetic variables, reducing concern for systematic bias. Second, the cross-sectional design precludes causal inference or conclusions about temporal trajectories; longitudinal studies are needed to determine whether these sex-specific sleep–memory associations predict cognitive decline or AD-related pathology. Third, although analyses focused on NREM physiology, future work should more comprehensively examine REM sleep, given emerging evidence linking REM disruption to AD risk.^80^ Accordingly, these findings should be considered hypothesis-generating rather than definitive evidence of sex differences. Replication in larger samples, including sufficient representation *APOE ε4* homozygotes, is needed to clarify genotype dose effects and enhance generalizability.

### 4.6. Conclusion

In summary, sex moderates associations between *APOE ε4* status, sleep physiology, and overnight memory retention, with the strongest sleep-memory relationships observed in this sample in female *ε4* carriers. These results suggest that sex-specific processes may shape how genetic risk for AD is expressed in brain function prior to clinical onset and highlights the importance of incorporating both sex and genotype into models of aging, sleep, and cognition. Further work is needed to clarify the neurobiological basis and translational relevance of these interactions. Given the modest sample size, findings should be interpreted cautiously. Overall, the results are consistent a model in which sex and genetic risk influence the functional relevance of distinct NREM sleep features for memory consolidation in the preclinical stage.

## Data Availability

All data produced in the present study are available upon reasonable request to the authors

## ^1^ Abbreviations

AD: Alzheimer’s disease
APOE ε4: apolipoprotein E epsilon 4
EEG: electroencephalography
hdEEG: high-density electroencephalography
NREM: non-rapid eye movement
REM: rapid eye movement
N2: stage 2 NREM sleep
N3: stage 3 NREM sleep.

## Acknowledgments

We thank the participants and staff of the Wisconsin Alzheimer’s Disease Research Center (ADRC) and Wisconsin Sleep for their contributions to this study.

## Consent Statement

All participants provided written informed consent prior to participation. Study procedures were approved by the Institutional Review Board at the University of Wisconsin-Madison.

